# Evaluating the Safety Profile of the CoronaVac in Adult and Elderly populations: A Phase IV Prospective Observational Study in Brazil

**DOI:** 10.1101/2023.08.19.23294316

**Authors:** Monica Akissue de Camargo Teixeira Cintra, Eder Gatti Fernandes, Vanessa Infante, Ana Paula Loch, Lucas Ragiotto, Patrícia Emília Braga, Maria da Graça Salomão, Maria Beatriz Bastos Lucchesi, Mayra Martho Moura de Oliveira, Vera Lúcia Gattás, Anderson Soares da Silva, Paulo José Fortes Villas Boas, Marta Heloisa Lopes, José Moreira, Fernanda Castro Boulos, CFV-01-IB study group

**Affiliations:** Clinical Trials and Pharmacovigilance Center, Instituto Butantan, São Paulo, Brazil; Centro de Saúde Escola da Faculdade de Medicina de Ribeirão Preto da Universidade de São Paulo (HCFMRP-USP) Dr. Joel Domingos Machado.; Centro de Saúde Escola da Faculdade de Medicina de Botucatu – Unesp.; Centro de Referência de Imunobiológicos Especiais Hospital das Clínicas da Faculdade de Medicina da Universidade de São Paulo (CRIE-HCFMUSP).

**Keywords:** CoronaVac, Adverse Events Following Immunization, Covid-19 vaccine, Adult vaccination, Active Pharmacovigilance, Phase IV trial, Covid-19

## Abstract

**Objective:** This Phase IV prospective observational study aimed to evaluate the safety and monitor adverse events following immunization (AEFI) associated with CoronaVac, an inactivated SARS-CoV-2 vaccine, in Brazilian adult (18-59 years) and elderly (≥60 years) populations.

**Methods:** Participants (n=538; 487 adults and 51 elderly) were enrolled from three public health services in São Paulo, Brazil. A two-dose vaccination regimen, administered 14 days apart, was used. The study assessed Adverse Reactions (AR) necessitating medical attention within seven days post-vaccination dose and monitored AEFI for a period of 42 days. Safety was monitored through a review of participant diary cards, telephone contacts, and on-site study visits.

**Results:** Among adults, the most frequently reported local AR after the first and second dose was pain (256[52.6%] and 129 [29.5], respectively), while the most common systemic reaction was a headache (158[34.5%] and 51 [11.6%], respectively). Most local and systemic solicited ARs were of Grade 1 or 2, with these reactions being more prevalent in adults after the first dose. One serious adverse event possibly related to the vaccine was reported among adults, but there were no fatalities. Nine adult participants experienced adverse events of special interest, which included five cases of Covid-19.

**Conclusion:** CoronaVac demonstrated safety and tolerability in the observed population. Ongoing post-marketing surveillance is crucial for the identification of rare adverse events and further affirmation of the vaccine’s safety profile.

## Introduction

The swift development of SARS-CoV-2 vaccines and their subsequent rapid market authorization process in response to the Covid-19 pandemic has underscore the importance of pharmacovigilance. This practice manages the substantial number of reported suspected adverse events following immunization (AEFI) and its implications for the global vaccination campaign [1].

Instituto Butantan (IB), a Brazilian public biomedical research and manufacturing institution, has played a pivotal role in Covid-19 governance in Brazil. In 2020, IB initiated a phase 3 clinical trial to evaluate efficacy and safety of CoronaVac in first-line healthcare professionals in Brazil [2]. In January 17^th^, CoronaVac was granted Emergency Use Authorization (EUA) by the National Health Surveillance Agency (ANVISA).

However, the large participants samples in a phase 3 trial have limitations in detecting rare adverse events. Routine pharmacovigilance activities typically depend on spontaneous reporting, which highlights the crucial role of post-marketing safety surveillance in prioritizing vaccine rollout and equitable access. Following ANVISA recommendations, IB undertook a post-market approval safety evaluation to characterize AEFI associated with CoronaVac in adults and the elderly during the vaccination campaign in Brazil.

## Methods

### Study design and setting

This phase IV prospective observational study monitored the safety of CoronaVac after a minimum of 1-dose administration. The study, registered on clinicaltrials.org prior to data acquisition (NCT04845048), was conducted from May 2021 to January 2022 across three public health services, in São Paulo state, Brazil. The study commenced after ANVISA issued an EUA for CoronaVac on January 17^th^, 2021. Ethical approval was obtained from the coordinating study site (University of São Paulo, protocol number: 4.684.670) and the collaborating study sites - Centro de Referência de Imunobiológicos Especiais/Hospital das Clínicas da Faculdade de Medicina da Universidade de São Paulo (CRIE-HCFMUSP), Centro de Saúde Escola da Faculdade de Medicina de Ribeirão Preto da Universidade de São Paulo (HCFMRP-USP), and Centro de Saúde Escola da Faculdade de Medicina de Botucatu – Unesp. Written informed consent was obtained from all study participants according to local IRBs. We reported as per the Strengthening the Reporting of Observational Studies in Epidemiology (STROBE) guideline (refer to Supplementary appendix).

### Participants

Eligible participants for the CoronaVac vaccine, aged 18-59 years-old (adults) and ≥ 60 years (elderly), were recruited based on the criteria of the national immunization program in Brazil during the study [3]. Potential participants were invited to join the study, with participation contingent on signing an informed consent form and agreement to adhere to study procedures. Individuals with a history of allergic reactions or anaphylaxis to previous immunizations, allergy to any CoronaVac component, fever (axillary temperature ≥ 37.8° C) within 72 hours prior to vaccination, or unavailable throughout the study period (42 days or 60 days in case of women with childbearing potential) were considered ineligible.

### Primary and secondary endpoints

The primary safety endpoint entailed the characterization of adverse reactions requiring medical care and assistance within 7 days post-administration of any of the two CoronaVac doses (14 days apart). The secondary endpoints encompassed: 1) characterization of AEFI requiring medical attention within 42 days post-administration of any dose, considering incidence, causality, and seriousness; 2) characterization of adverse reactions within 30 minutes post-administration of any dose, considering incidence and seriousness, 3) characterization of serious adverse event (SAE) within a 42-day period post-administration of any dose of the vaccine, stratified by age group, and 4) profiling of adverse event of special interest (AESI) within a 42-day period post-administration of any dose of the vaccine, stratified by age group. Adverse events were categorized on dose incidence, causality, severity, and age group. The exploratory endpoints included documenting adverse events among pregnant women and their exposed infants.

### Study procedures for active surveillance

The study entailed two visits to the clinical centers (V1 -initial screening and first dose vaccination; V2 - second dose vaccination and surveillance assessment through participant diary card review, and seven telephone contacts (C1 to C7 - for active AE surveillance). If eligible, participants received the first vaccine dose and remaining under observation at the clinical center for 30 minutes for immediate hypersensitivity AEFI assessment and solicited AEFI recording (V1 and V2). Clinical center staff also gathered information concerning demographics (sex, age, and race), medical history, concomitant medication, ongoing pregnancies and infants (where applicable), vaccination site (right arm/left arm/other), administration route (intramuscular/other) and volume (0.5mL/other), vaccine batch, manufacturing date and expiration date.

After vaccination, each participant was evaluated by study site staff for immediate occurrence of solicited and unsolicited AEFI. Before departing, participants received a diary card for solicited and unsolicited adverse events up to 7 days post-vaccination, a device for measuring local site AEFI, a thermometer, and instructions for use. Participants were further instructed to return to the study site for a second visit (V1 + 14 days tolerance), respond to scheduled telephone calls [C1 (V1 + 3-5 days) and C2 (V1 + 7-9 days)], and promptly notify study staff in the event of hospitalization and/or SAE. Participants were also provided instructions on how to record AEFIs and assess the severity/intensity of the AEFIs using a simple scoring system. AEFIs occurring during pregnancy or related to newborns were documented, if applicable. Information regarding AEFIs recorded in participant diary card and solicited AEFIs were collected by the sites during subsequent telephone contacts [C3 (V2 + 3-5 days), C4 (V2 + 7-9 days), C5 (V2 + 14-16 days), C6 (V2 + 42-45 days) and C7 – end of study (V2 + 60-63 days)]. During the second visit (V1 + 14 - 31 days), participants underwent physical examination and concomitant medication review. Possible pregnancy information was gathered, if applicable. Data recorded in the participant dairy cards were scrutinized by site center medical staff.

### Safety assessment

In this context, an adverse event was defined as any undesirable medical occurrence that affected a vaccinated participant, regardless of its causal relationship to the vaccine. Consequently, an adverse event could comprise any unfavorable and unintended sign, symptom, or disease (including an abnormal laboratory findings), temporally associated with the vaccination. All adverse events were encoded and categorized in accordance with the Medical Dictionary for Regulatory Activities (MedDRA, version 24.0) [3]. A Serious Adverse Event (SAE) was characterized as any adverse event resulting in outcomes such as death, a life-threatening condition, the necessity for hospitalization or an extension of an existing hospitalization, significant or persistent incapacitation, congenital anomaly, any suspected transmission of an infectious agent through a medicinal product, or any clinically significant adverse event. An Adverse Reaction (AR) was defined as any AE with a plausible causal relationship with the vaccine, as determined by the adapted classification of the Uppsala Monitoring Centre (UMC) of the World Health Organization (WHO) [4][5]. An adverse event of special interest was a predefined, medically significant event that might potentially be causally associated with a vaccine product, warranting close monitoring and confirmation via additional specific studies [6].

The severity of an AE was classified from grade one to four according to the Toxicity Grading Scale for Healthy Adult and Adolescent Volunteers Enrolled in Preventive Vaccine Clinical Trials of the US Food and Drug Administration (FDA) [7]. The study staff was instructed to incorporate the following information in their AE evaluations: the classification of seriousness; the classification of severity [4], causality assessment, and adverse event outcome (Supplementary information).

The definitions and classifications of adverse events were in accordance with the recommendations stipulated by the Good Clinical Practices (GCP) [8], the Guideline on Clinical Safety Data Management from the International Conference on Harmonization (ICH) [5], and the Brighton Collaboratiońs guidance on safety data collection for COVID-19 vaccine safety [6]. In instances where case definitions were not provided by Brighton Collaboration sources, the Common Terminology Criteria for Adverse Events (CTCAE) issued by the US Department of Health and Human Services [9] were utilized as reference.

#### Study vaccine

The study vaccine is an inactivated whole-virion vaccine with aluminum hydroxide serving as the adjuvant, prepared with a novel coronavirus strain (CZ02 strain) inoculated in African green monkey kidney cells (Vero cells) [10]. The inactivation process involved adding β-propiolactone to the virus harvest fluid at a ratio of 1:4000 and inactivating at 2-8° C for 12-24 h. A single dose of the vaccine contains 3 μg of SARS-CoV-2 virion in a 0.5 mL aqueous suspension for injection with 0.45 mg/mL of aluminum. The vaccine was administered in two doses, delivered 14 days apart, intramuscularly in the deltoid muscle. The vaccine is free of preservatives and the final packaging includes a single-dose vial with an extractable volume of 0.5 mL, a vial with two doses of 0.5 mL each, or a multi-dose vial containing 5 mL, with each vaccine dose corresponding to 0. 5 ml.

### Sample size estimation

The sample size was estimated following the recommendation of the Guide to Clinical Evaluations of New Vaccines (EMEA/CHMP/VWP/164653/2005) [11], suggesting that at least 100 participants from each vaccination target group should have AEFI monitored. Thus, a sample of 900 vaccinated participants (600 adults and 300 elderly) was estimated for the evaluation of solicited and unsolicited AEFI through active surveillance.

This sample size was based on the formula presented below, assuming the observation of an AE at an established minimum frequency equal to M (maximum risk). N = ln(α)/ln(1-M) Type I error parameter (α) adopted was 5% (two-tailed) and N for elderly=300, to achieve AE frequency detection of approximately 1% (maximum risk of 1 to 100), whereas N for adults= 600 achieves AE frequency at a maximum risk of 1 in 200.

### Statistical analysis

Descriptive statistical data analysis of AE and AR was performed, stratified by study groups (younger and older adults) and dose vaccination [12]. Results are presented as the frequency of occurrence (absolute and percentage values) for qualitative variables and as dispersion measures (quartiles) for quantitative variables. Missing data were handled with the list-wise deletion method. The statistical analyses were conducted using Stata (Stata-Corp LP, College Station, Texas USA), version 13.0, for Windows.

## Results

### Participants

Figure 1 presents the study participantś flowchart, segmented to age groups. The study incorporated a total of 538 participants, where young adults represented 90.5% (487/538) of the sample and older adults constituted 9.5% (51/538). Table 1 provides a description of the participants’ baseline socio-demographic characteristics. The median age for young adults was 30.5 years, and for the oldest adults, its was 65.3 years. The majority of participants were female (57.1%) and identified as white (59.3%).

**Figure 1.**
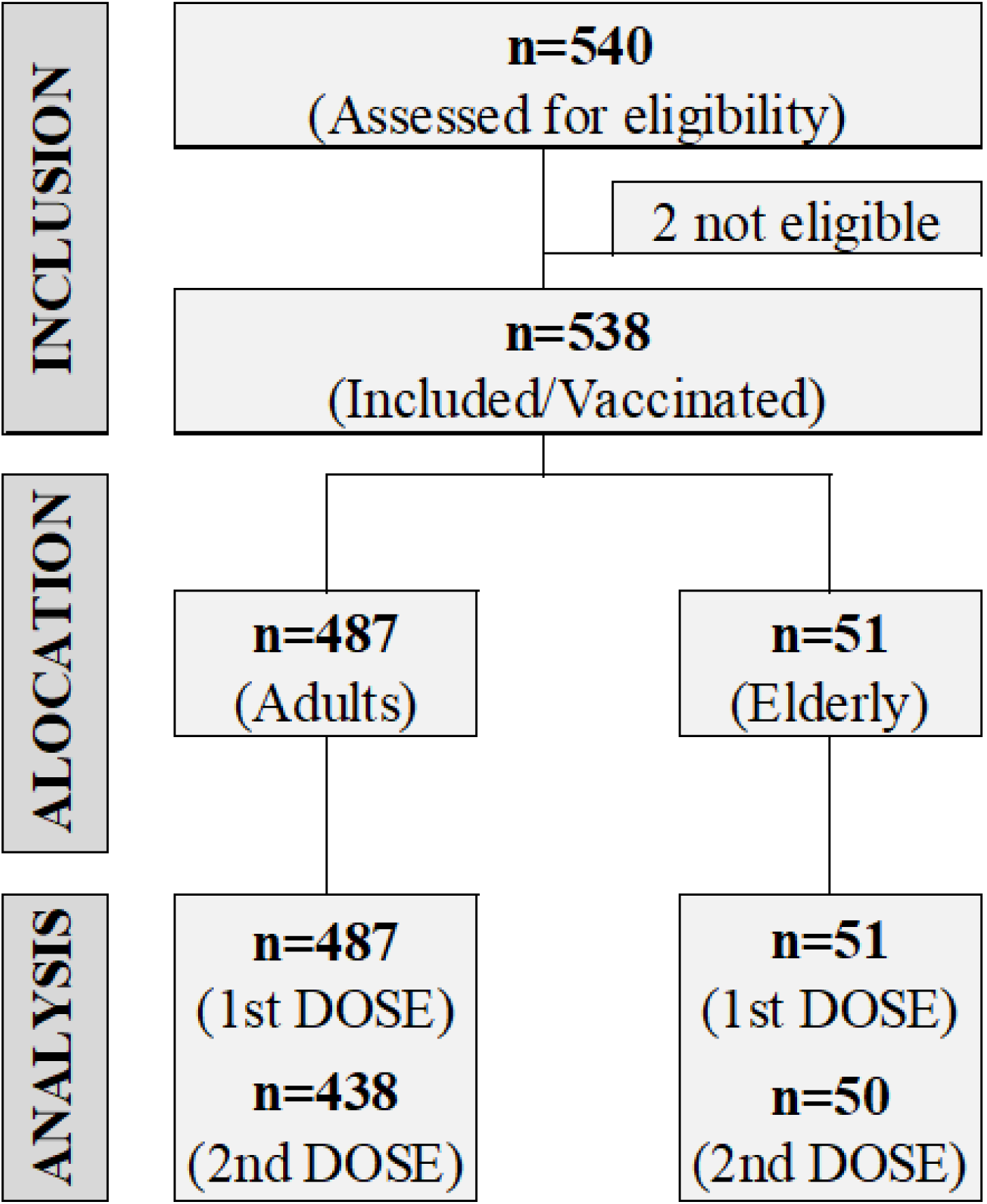
Study flowchart: screening, enrollment, and vaccination.

**Table 1.**
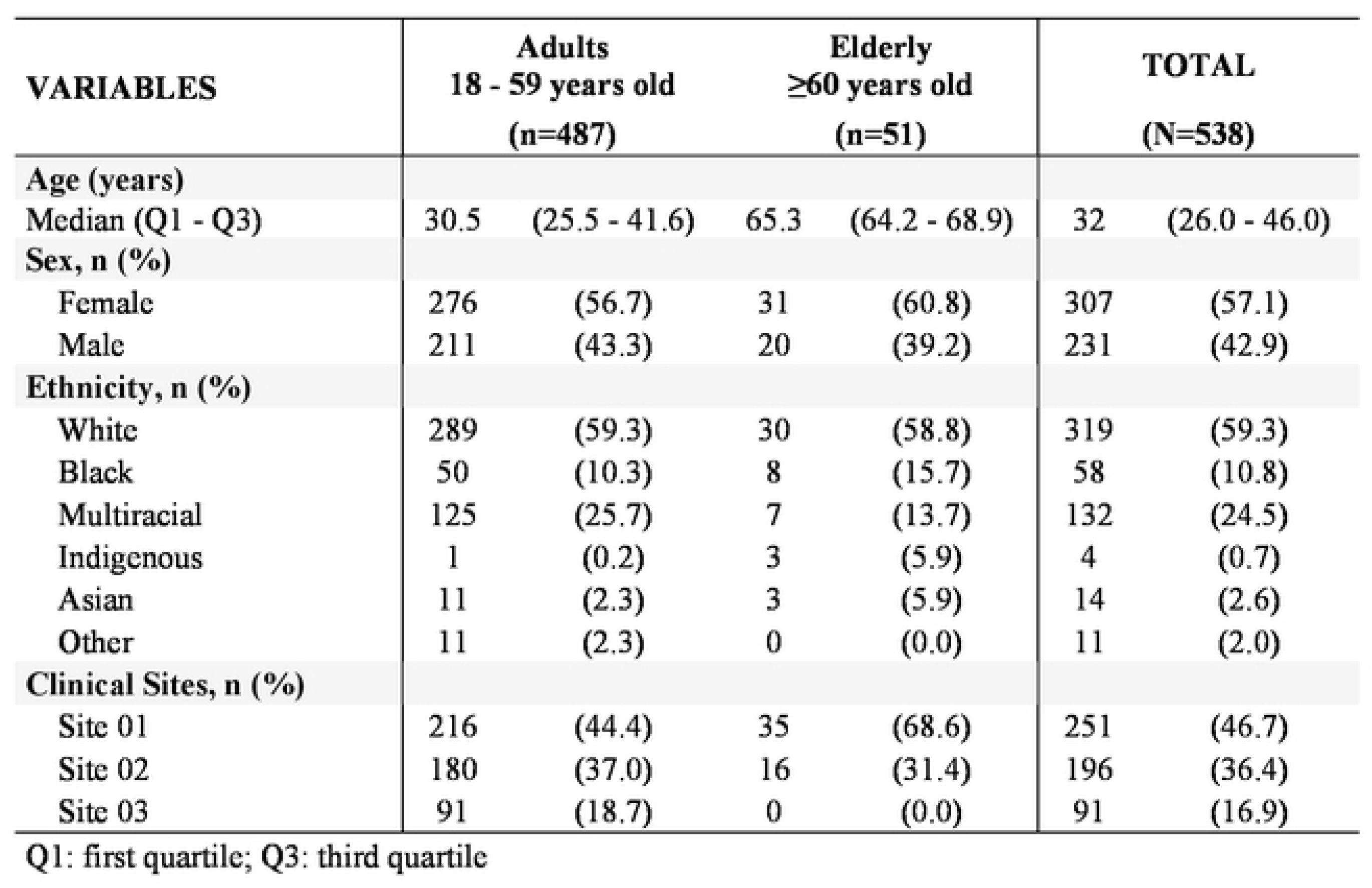
Demographic characteristics of all participants by study group.

The median occurrence of AE and AR per participant was three following the first dose, and varied from one to three following the second-dose., across the entire population (Table 2). Both AE and AR were more frequent in the 18-59-year-old group after the first dose (V1). Only one SAE was reported as possibly related to vaccination, with no registered cases of death or permanent impairment. The AE and AR were more frequent after V1 in the 18–59-year-old group.

**Table 2.** Frequency of Adverse Events and Adverse Reactions after Coronavac vaccination (first and second dose) bystudy group.

| STUDY GROUP | 1st DOSE |  |  |  | 2nd DOSE |  |  |  |
| --- | --- | --- | --- | --- | --- | --- | --- | --- |
|  | AE<br>n | Participants with AE<br>n/n<sub>total</sub> (%) | AE per participant |  | AE<br>n | Participants with AE<br>n/n<sub>total</sub> (%) | AE per participant |  |
|  |  |  | median | (Q<sub>1</sub>-Q<sub>3</sub>) |  |  | median | (Q<sub>1</sub>-Q<sub>3</sub>) |
| Adults (18 - 59 years) | 1569 | 392/487 (80.5) | 3 | (2 - 5) | 554 | 218/438 (49.8) | 2 | (1 - 3) |
| Elderly (≥60 years) | 136 | 30/51 (58.8) | 3 | (1 - 6) | 94 | 28/50 (56.0) | 3 | (1 - 5) |
| **TOTAL** | 1705 | 422/538 (78.4) | 3 | (2 - 5) | 648 | 246/488 (50.4) | 2 | (1 - 3) |
| STUDY GROUP | 1st DOSE |  |  |  | 2nd DOSE |  |  |  |
|  | AR<br>n | Participants with AR<br>n/n<sub>total</sub> (%) | AR per participant |  | AR<br>n | Participants with AR<br>n/n<sub>total</sub> (%) | AR per participant |  |
|  |  |  | median | (Q<sub>1</sub>-Q<sub>3</sub>) |  |  | median | (Q<sub>1</sub>-Q<sub>3</sub>) |
| Adults (18 - 59 years) | 1288 | 374/487 (76.8) | 3 | (1 - 5) | 438 | 194/438 (44.3) | 2 | (1 - 3) |
| Elderly (≥60 years) | 84 | 23/51 (45.1) | 3 | (1 - 4) | 53 | 22/50 (44.0) | 1 | (1 - 4) |
| **TOTAL** | 1372 | 397/538 (73.8) | 3 | (1 - 5) | 491 | 216/488 (44.3) | 2 | (1 - 3) |

Q1: first quartile; Q3: third quartile
AE: Adverse Event
AR: Adverse Reaction

### Primary endpoint

For adults, considering the primary endpoints, 76.0% experienced solicited AR after the first dose, and 42.5% after the second dose, within a 7-day post-vaccination period (Table 3). The most common local AR after both doses was pain at the administration site (256 [52.6%] and 129 [29.5%], respectively). The majority of local and systemic solicited AR were classified as Grade 1 or Grade 2 (Table S1). Thirty-five cases (3.0%) were classified as Grade 3 after the first dose, while 10 cases (2.6%) after the second. Only three ARs were classified as Grade 4 (1 headache, 1 fatigue and 1 myalgia), all following the first dose.

**Table 3.** Frequency of solicited adverse reactions (AR}7 days post vaccination in adults (18 - 59 years).

| Solicited Adverse Reactions (AR) | 1st DOSE |  |  | 2nd DOSE |  |  |
| --- | --- | --- | --- | --- | --- | --- |
|  | AR Adults<br>(n=487) |  | AR Total | AR Adults<br>(n=438) |  | AR Total |
|  | n | % | n | n | % | n |
| Local |  |  |  |  |  |  |
| Pain | 256 | 52.6 | 263 | 129 | 29.5 | 132 |
| Erythema | 9 | 1.8 | 9 | 10 | 2.3 | 11 |
| Swelling | 15 | 3.1 | 15 | 3 | 0.7 | 4 |
| Induration | 10 | 2.1 | 11 | 3 | 0.7 | 3 |
| Pruritus | 15 | 3.1 | 15 | 4 | 0.9 | 4 |
| Tenderness | 166 | 34.1 | 169 | 59 | 13.5 | 59 |
| Systemic |  |  |  |  |  |  |
| Decreased appetite | 20 | 4.1 | 20 | 6 | 1.4 | 7 |
| Arthralgia | 40 | 8.2 | 44 | 11 | 2.5 | 11 |
| Chills | 35 | 7.2 | 37 | 9 | 2.1 | 9 |
| Headache | 168 | 34.5 | 191 | 51 | 11.6 | 54 |
| Diarrhea | 36 | 7.4 | 38 | 14 | 3.2 | 15 |
| Rash | 8 | 1.6 | 8 | 1 | 0.2 | 1 |
| Fatigue | 107 | 22.0 | 127 | 23 | 5.3 | 25 |
| Fever (Pyrexia) | 14 | 2.9 | 14 | 5 | 1.1 | 5 |
| Hypersensitivity (Allergic Reaction) | 2 | 0.4 | 2 | 0 | 0.0 | 0 |
| Myalgia | 69 | 14.2 | 72 | 21 | 4.8 | 22 |
| Nausea | 39 | 8.0 | 43 | 16 | 3.7 | 17 |
| Pruritus Outside the Administration Site | 32 | 6.6 | 33 | 5 | 1.1 | 5 |
| Cough | 38 | 7.8 | 39 | 5 | 1.1 | 5 |
| Vomiting | 6 | 1.2 | 7 | 2 | 0.5 | 2 |
| TOTAL SOLICITED | 370 | 76.0 | 1157 | 186 | 42.5 | 391 |

For the elderly, considering the primary endpoints, 43.1% experienced solicited AR after the first dose, and 44.0 % after the second dose, within a 7-day post-vaccination period (Table 4). The most common local AR was pain at the administration-site (9 [17.7%] and 11 [22.0%] after first and second dose, respectively) and the most frequent systemic AR one was headache (9 [17.7%] and 10 [20.0%] after first and second dose, respectively). The majority of local and systemic solicited AR were classified as Grade 1 or Grade 2 (Table S2). Only one episode of myalgia was classified as Grade 3, no local and systemic solicited AR were classified as Grade 4.

**Table 4.**
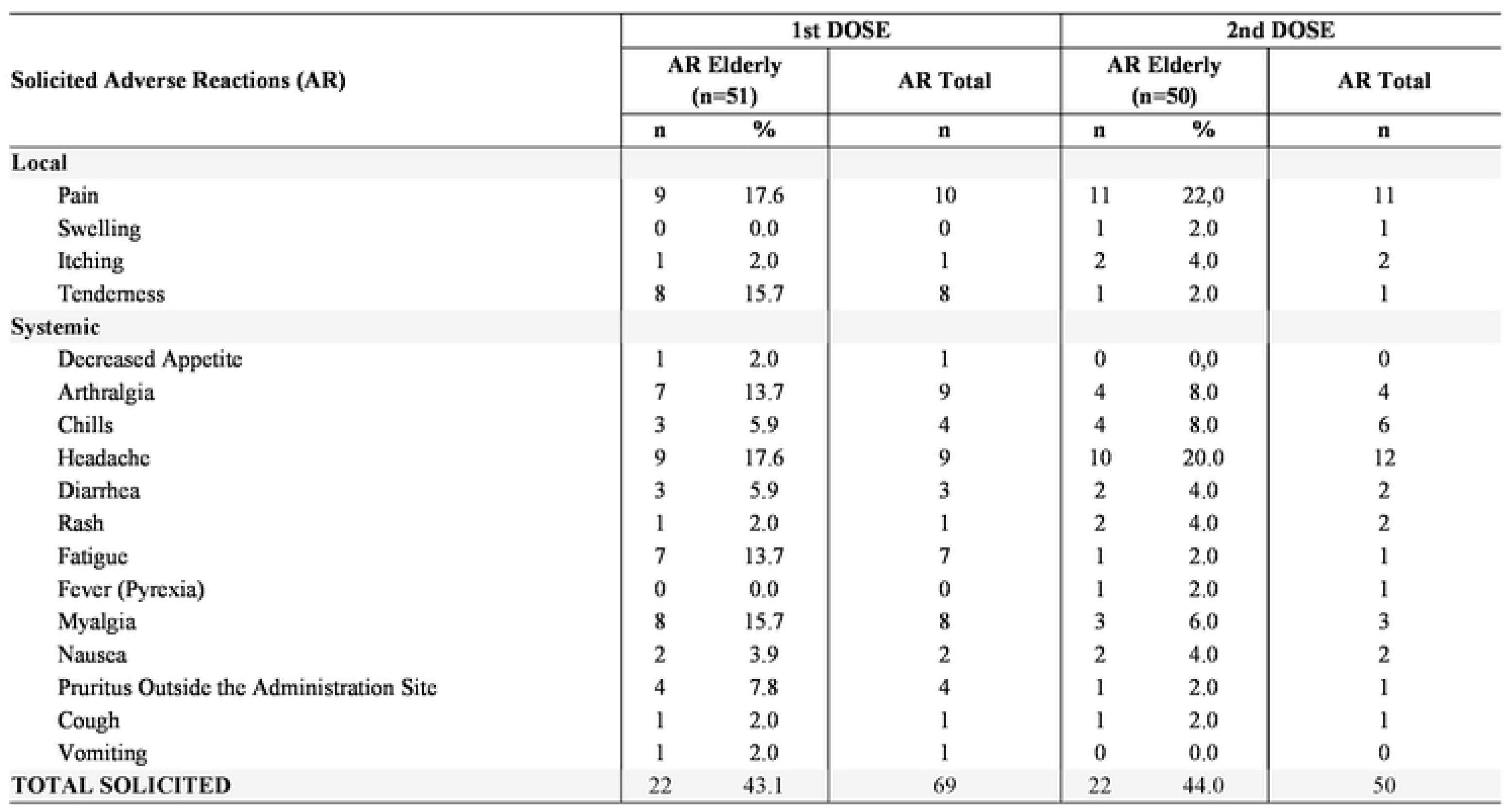
Frequency of solicited adverse reactions (AR}7 days post vaccination in elderly (:::60 years).

Frequency and severity of solicited and unsolicited AR within 7-days post-vaccination for adults and elderly are available in the Supplementary material (Table S1 and Table S2).

### Secondary endpoint

Table 5 displays the frequency of adults and elderly experiencing solicited AR 30 minutes post each vaccination. The solicited AR were more common after the first dose among adults (129 [26.5%] vs 84 [19.2%]). Data suggests that solicited AR were less frequent among the elderly, with no noticeable difference between the first and the second dose (6 [11.8%] and 6 [12.0%], respectively). The frequency and intensity of unsolicited AR 30 minutes post-vaccination in adults and elderly can be found in the Supplementary material (Table S3).

**Table 5.** Frequency of Adverse Reactions (AR) within 30 minutes post vaccination in adults (18 - 59 years) and elderly (l:60 years).

| Solicited Adverse Reactions (AR) | 1st DOSE |  |  | 2nd DOSE |  |  | 1st DOSE |  |  | 2nd DOSE |  |  |
| --- | --- | --- | --- | --- | --- | --- | --- | --- | --- | --- | --- | --- |
|  | AR Adults<br>(n=487) |  | AR Total | AR Adults<br>(n=438) |  | AR Total | AR<br>Elderly<br>(n=51) |  | AR Total | AR<br>Elderly<br>(n=50) |  | AR Total |
|  | n | % | n | n | % | n | n | % | n | n | % | n |
| **Local** |  |  |  |  |  |  |  |  |  |  |  |  |
| Pain | 85 | 17.5 | 85 | 52 | 11.9 | 52 | 4 | 7.8 | 4 | 6 | 12.0 | 6 |
| Erythema | 2 | 0.4 | 2 | 2 | 0.5 | 2 | 0 | 0.0 | 0 | 0 | 0.0 | 0 |
| Induration | 5 | 1.0 | 5 | 3 | 0.7 | 3 | 0 | 0.0 | 0 | 0 | 0.0 | 0 |
| Itching | 1 | 0.2 | 1 | 1 | 0.2 | 1 | 0 | 0.0 | 0 | 1 | 2.0 | 1 |
| Tenderness | 48 | 9.9 | 48 | 32 | 7.3 | 32 | 2 | 3.9 | 2 | 1 | 2.0 | 1 |
| **Systemic** |  |  |  |  |  |  |  |  |  |  |  |  |
| Arthralgia | 0 | 0.0 | 0 | 1 | 0.2 | 1 | 0 | 0.0 | 0 | 1 | 2.0 | 1 |
| Headache | 9 | 1.8 | 9 | 4 | 0.9 | 4 | 1 | 2.0 | 1 | 0 | 0.0 | 0 |
| Fatigue | 3 | 0.6 | 3 | 3 | 0.7 | 3 | 0 | 0.0 | 0 | 0 | 0.0 | 0 |
| Myalgia | 3 | 0.6 | 3 | 1 | 0.2 | 1 | 0 | 0.0 | 0 | 0 | 0.0 | 0 |
| Nausea | 3 | 0.6 | 3 | 2 | 0.5 | 2 | 0 | 0.0 | 0 | 0 | 0.0 | 0 |
| Cough | 1 | 0.2 | 1 | 1 | 0.2 | 1 | 0 | 0.0 | 0 | 0 | 0.0 | 0 |
| Vomiting | 0 | 0.0 | 0 | 0 | 0.0 | 0 | 0 | 0.0 | 0 | 0 | 0.0 | 0 |
| **TOTAL SOLICITED** | 129 | 26.5 | 160 | 84 | 19.2 | 102 | 6 | 11.8 | 7 | 6 | 12.0 | 9 |

The frequency of AR that required medical intervention and the SAE 42-days post vaccination are outlined in Table S4. There were no SAE reports in the elderly group. Only one SAE (migratory polyarthralgia) was assessed by the Investigator and considered possibly related to the vaccine. All participants that presented any AR that required medical intervention or SAE had fully recovered.

Nine adult participants experienced AESI. These included five cases of Covid-19, two cases of arthritis, one case of tachycardia, and one case of acute renal failure. The frequency, causality assessment and intensity of the AESI can be found at Supplementary material (Table S5).

### Exploratory endpoints

Among pregnant participants (n=2), seven non-serious AR were reported. All these cases recovered. Detailed information about the AE in pregnant women and infants can be found in Supplementary material (Table S6).

## DISCUSSION

The expedited development and EUA of Covid-19 vaccines have been instrumental in mitigating the global impact of the SARS-CoV-2 pandemic, especially during its acute phase [13]. Among these vaccines, CoronaVac, developed by Sinovac Biotech and later transferred to IB, has been extensively utilized in numerous countries. However, upon EUA, safety data for CoronaVac, as with all Covid-19 vaccines authorized during this public health crisis, were limited, thus necessitating rigorous post-vaccination real-world safety data.

The ANVISA recognizing this knowledge gap commissioned a comprehensive post-authorization safety study on CoronaVac in adults and elderly populations. The aim was to document its safety profile, particularly focusing on the identification and characterization of AEFI among adults and the elderly populations. The real-world nature of this study offered the opportunity to evaluate the vaccinés safety profile in a broader and more diverse population than those included in clinical trials. Our findings suggest that most AE and AR following vaccination were mild to moderate in severity and resolved spontaneously. The frequency of AR, solicited and unsolicited, was greater in younger adults than in elderly, especially after the first dose. Only one SAE was reported, suggesting a satisfactory safety profile of the vaccine.

Brazilian national passive surveillance data reported a 6.6% rate of SAE following CoronaVac [14], whereas our study identified a significantly lower rate of 0.6%. Unlike passive pharmacovigilance, which relies on voluntary spontaneous reporting of AE, active pharmacovigilance facilitates the procurement of more detailed safety information. The passive surveillance is more sensitive to SAE, but the data quality is weak [15][16]. A strength of our research is the active surveillance of AEFI by follow-up phone calls after each dose. This procedure made possible to capture more detailed mild and moderate events, which are typically underreported in passive surveillance systems [15]. Another strength of this study is the low dropout rate, which reinforces the good tolerability of the vaccine. Additionally, the participants presented different risk levels of exposure to Covid-19.

Our study has limitations. Sample size did not reach the previously calculated number of participants because the public vaccination campaign started 3 months earlier than the protocol approval, including other vaccines rather than CoronaVac. Due to this difficulty to include unvaccinated subjects, and therefore, the reduction in sample size, the minimum of AE detection power for adults was M=1/165≈0.6% whereas for elderly was M=1/18 ≈5.7%. The demographic distribution of the sample population, which had a considerably larger representation of younger adults than older ones might potentially introduce some bias in the results and explain the observed higher vaccine reactogenicity in the younger adults compared to older ones. After CoronaVaćs EUA extension to the pediatric population in January 20^th^, 2022, a protocol amendment was made to include children and adolescents, enabling safety information collection for this group. Albeit the above sample size limitations, our data are consistent with other studies that suggest CoronaVac are well tolerated with a relatively lower frequency of AE regardless of age [10] [17] [18].

Our findings showed that the majority of ARs were mild to moderate [Grade 0 and Grade 1: (962/1157 - 83.1%)], which coincides with previous reports evaluating this vaccine [10] [17] [18]. Most AE were low grade and resolved spontaneously, supporting the findings reported by Tanriover et al. [10] and Wu et al. [19] in Turkey and China, respectively. Most of the solicited AR reported by the participants lasted less than a day, differently from data in Xia et al. [20] as well as Zhang et al. [21] studies which reported adverse reactions lasting for up to 72h after the complete schedule of vaccine administration in healthy adults. Most common adverse events reported in our study, such as local pain, headache, and myalgia, were also observed in other studies evaluating CoronaVac and are listed in the vaccine package insert. Regarding the most frequent local and systemic adverse reactions by inactivated virus Covid-19 vaccine, Ling et al. [22] reported pain, swelling, and fever at the injection site, whereas Chen et al. [23] reported pain, swelling and redness at the injection site. Systemic AEFI such as headache (34.5%) and fatigue (21.97%) predominated in our study. Results from randomized trials in Indonesia have listed headache and malaise as the most significant systemic AEFI [24]. Indonesian results pointed out gender related AEFI after CoronaVac immunization in which females presented higher frequency of AEFI than male participants.

Our study adds to the growing body of evidence on the safety of SARS-CoV-2 vaccines, particularly CoronaVac. It equips clinicians with pertinent information for patient counseling about potential CoronaVac side effects and provides policymakers with crucial data to inform vaccination strategies and campaigns. In fact, our data helped consolidate the safety profile of CoronaVac in the Braziliańs Covid-19 vaccine response plan.

In conclusion, our data substantiate the overall safety of the CoronaVac in adults and elderly individuals, with side effects being generally mild and transient. However, continuous monitoring for rare AE remains crucial. These findings should bolster confidence in CoronaVac among clinicians, policymakers, and the public, while encouraging further research on Covid-19 vaccines in various demographic and epidemiological settings.

## Acknowledgments

We thank the study volunteers. The studýs conception, design, conduct, analysis, and interpretation, as well as the drafting and submission of this manuscript, involved the IB staff. The corresponding author had full access to the study data and had final responsibility for the decision to submit for publication.

## Declarations

## Consent to participate

All participants gave informed consent to participate in this study, where publication was identified as one form of dissemination. Information and Study Consent Forms available upon request.

## Ethics approval

Approval for this research was granted by the Ethical Committee of the University of São Paulo (protocol number: 4.684.670).

## Data availability

All data are available in the text and in the supplementary appendix for consultation.

## Supporting information captions

S1 Table. Frequency and severity of adverse reactions, solicited (local and systemic) and unsolicited, occurring up to 7 days after administration of each vaccine dose in adults (18 to 59 years), according to severity

S2 Table. Frequency and severity of adverse reactions, solicited (local and systemic) and unsolicited, occurring up to 7 days after administration of each vaccine dose in elderly (≥60 years), according to severity

S3 Table. Frequency of adverse reactions, solicited (local and systemic) and unsolicited, occurring within 30 minutes after administration of each vaccine dose in a) adults (18-59 yeas) and b) elderly (≥60 years), according to severity

S4 Table. Frequency of solicited (local and systemic) and unsolicited adverse reactions which required medical attention, occurring at any time within 42 days after administration of each vaccine dose in a) adults (18-59 years) and b) elderly (≥60 years), according to severity

S5 Table. Serious adverse events and adverse events of special interest according to the administration dose regarding description (MedRA code), causality, severity, predictability and outcomes

S6 Table. Adverse event details observed in pregnant women and newborn according to the administration dose regarding description code (MedRa), causality, severity, predictability and outcomes

